# Metabolomic and genomic prediction of common diseases in 477,706 participants in three national biobanks

**DOI:** 10.1101/2023.06.09.23291213

**Authors:** Nightingale Health Biobank Collaborative Group, Jeffrey C. Barrett, Tõnu Esko, Krista Fischer, Luke Jostins-Dean, Pekka Jousilahti, Heli Julkunen, Tuija Jääskeläinen, Nurlan Kerimov, Sini Kerminen, Anastassia Kolde, Harri Koskela, Jaanika Kronberg, Sara N. Lundgren, Annamari Lundqvist, Valtteri Mäkelä, Kristian Nybo, Markus Perola, Veikko Salomaa, Kirsten Schut, Maiju Soikkeli, Pasi Soininen, Mika Tiainen, Taavi Tillmann, Peter Würtz, the Estonian Biobank Research Team

## Abstract

Identifying individuals at high risk of chronic diseases via easily measured biomarkers could improve public health efforts to prevent avoidable illness and death. Here we present nuclear magnetic resonance blood metabolomics from half a million samples from three national biobanks. We built metabolomic risk scores that identify a high-risk group for each of 12 diseases that cause the most morbidity in high-income countries and show consistent cross-biobank replication of the relative risk of disease for these groups. We show that these metabolomic risk scores are more strongly associated with future disease onset than polygenic scores for most of these diseases. In a subset of 18,000 individuals with metabolomic biomarkers measured at two time points we show that people whose scores change have dramatically different future risk of disease, suggesting that repeat measurements capture the benefits of lifestyle change. We show cross-biobank calibration of our scores. Since metabolomics can be measured from a standard blood sample, we propose such tests can be feasibly implemented today in preventative health programs.

**One-Sentence Summary:** Biomarkers from half a million blood samples identifies people at increased risk of chronic diseases and can be used for early detection today.

---

Life expectancy is increasing faster than healthy life expectancy, leading to individuals in high-income countries to live more years restricted by chronic diseases (*1, 2*). Healthcare systems face a cost crisis as they try to provide an increasingly wide range of transformative but expensive treatments to the older, sicker populations they care for (*3*). Better prevention, to complement new treatments, is essential to provide healthier lives and to financially sustain healthcare systems. The established approach to preventative health is: (i) identify affordable interventions without side effects that can reduce future disease incidence, (ii) develop risk prediction algorithms to identify the right individuals to target, and (iii) implement the risk prediction as widely as possible. For example, the NHS Health Check (*4*) is available to adults in the UK every five years, and uses lifestyle factors, family and clinical history, blood pressure, and a cholesterol test to identify individuals at high risk of cardiovascular disease who should adjust their lifestyle or begin taking cholesterol-lowering or blood pressure reducing medicine (*5*).

Risk stratification can be improved for cardiovascular disease and applied to a much wider range of diseases via ‘omic’ data types that have become mainstays of medical research but are not yet incorporated into healthcare. Most focus has been on polygenic scores (PGS) (*6, 7*), which can identify individuals at elevated risk for multiple diseases (*8*) only need one measurement, and offer complementary information to traditional risk factors (*9, 10*). Implementation of PGS has been limited by both resistance to broader clinical use of genetic information, and practical challenges to measuring a DNA sample and inserting the relevant information to health records. Metabolomic risk models, based on biomarkers measured in blood samples, for example via nuclear magnetic resonance spectroscopy (*11–13*), have also been shown to predict many common diseases (*14, 15*) including cardiovascular events, type 2 diabetes (*16*), and all-cause mortality (*17*). The metabolomic measurement needed for these scores can be generated from a standard blood sample, and includes many biomarkers already familiar in clinical practice, such as cholesterol, glucose, and creatinine. Furthermore, since the metabolomic risk scores may change in response to lifestyle and treatment (in contrast to PGS), they can be used both to identify high-risk individuals and to track changes in their risk profile. A few studies have suggested complementary value for genetics and metabolomics in cardiovascular disease and type 2 diabetes (*18, 19*), but the combined use of these omics-based risk predictors has not yet been evaluated at scale.

Here, we generated nuclear magnetic resonance metabolomic data in half a million samples from individuals with years of follow-up data on clinical outcomes to train and test risk prediction of the twelve leading causes of disability-adjusted life years (DALYs) in high-income countries. We investigate the relative prediction of metabolomic scores and PGS in different diseases and time scales, assess the value of multiple metabolomic time points, and discuss how such risk predictions could plausibly be used in real-world public health settings.

## Metabolomic risk prediction across top sources of morbidity

### Building risk models

We collaborated with the UK Biobank, Estonian Biobank, and Finnish THL Biobank to measure metabolomic biomarkers via nuclear magnetic resonance spectroscopy in blood samples provided at the time of enrolment from 477,706 individuals with linked comprehensive clinical data (**Table 1, Table S1**). All three biobanks contain adults from Northern European countries, with varying ascertainment, recruitment years, age ranges, and procedures for extracting outcomes from electronic health records (**Methods, Figure S1**).

**Table 1.** Basic characteristics of the participants in the three national biobanks. See Figure S1 for age and recruitment year histograms. *See Table S1 for characteristics by cohort.

|  | **UK Biobank** | **Estonian Biobank** | **THL Biobank\*** |
| --- | --- | --- | --- |
| **Number of participants** | 254,993 | 190,785 | 31,928 |
| **Age at blood sample**<br>**(median, [IQR])** | 58.0 [50.0-63.0] | 43.0 [31.0-56.0] | 50.0 [39.0-61.0] |
| **Females (%)** | 54.0 | 65.8 | 53.5 |
| **Body mass index**<br>**(kg/m<sup>2</sup>; median, [IQR])** | 26.7 [24.2-29.8] | 25.3 [22.4-29.0] | 26.2 [23.6-29.3] |
| **Smoking prevalence (%)** | 10.5 | 18.3 | 34.8 |
| **Cholesterol-lowering**<br>**medication (%)** | 17.1 | 10.1 | 10.6 |
| **Follow-up time**<br>**(median, [IQR])** | 11.8 [11.1-12.5] | 3.2 [2.9-3.7] | 13.8 [8.8-15.2] |
| **Recruitment period** | 2006-2010 | 2002-2021 | 1997-2012 |

We analysed 12 diseases causing the most morbidity in the WHO European region (excluding falls and back pain, **Fig. 1, Table S2**), causing more than one third of all disability adjusted life years. We trained cox proportional hazards models to predict incidence of each of these diseases in a random half of UK Biobank (128,288 individuals). We included age and sex in all models as fixed covariates and allowed the model to select (via LASSO with five-fold cross-validation) from among 36 metabolomic biomarkers that have been validated in Europe for use in an in vitro diagnostic medical device (**Methods**). We evaluated the performance of these models in the remaining 50% of UK Biobank, as well as the Estonian and Finnish THL biobanks. As we quantify the biomarkers in absolute concentration units (e.g. mmol/l), we can evaluated directly using the score weights estimated in the UK Biobank in the other two datasets, without normalizing the biomarkers within each study separately. This is distinct from common practice in other omics analyses, where within cohort normalization is essential (*20, 21*). **Figure S2** shows that we obtain highly similar results with these normalization steps, but we here present results without this step to better mimic predicting a new individual’s risk without additional information (e.g. batch corrections, or cohort means and variances).

**Fig. 1.**
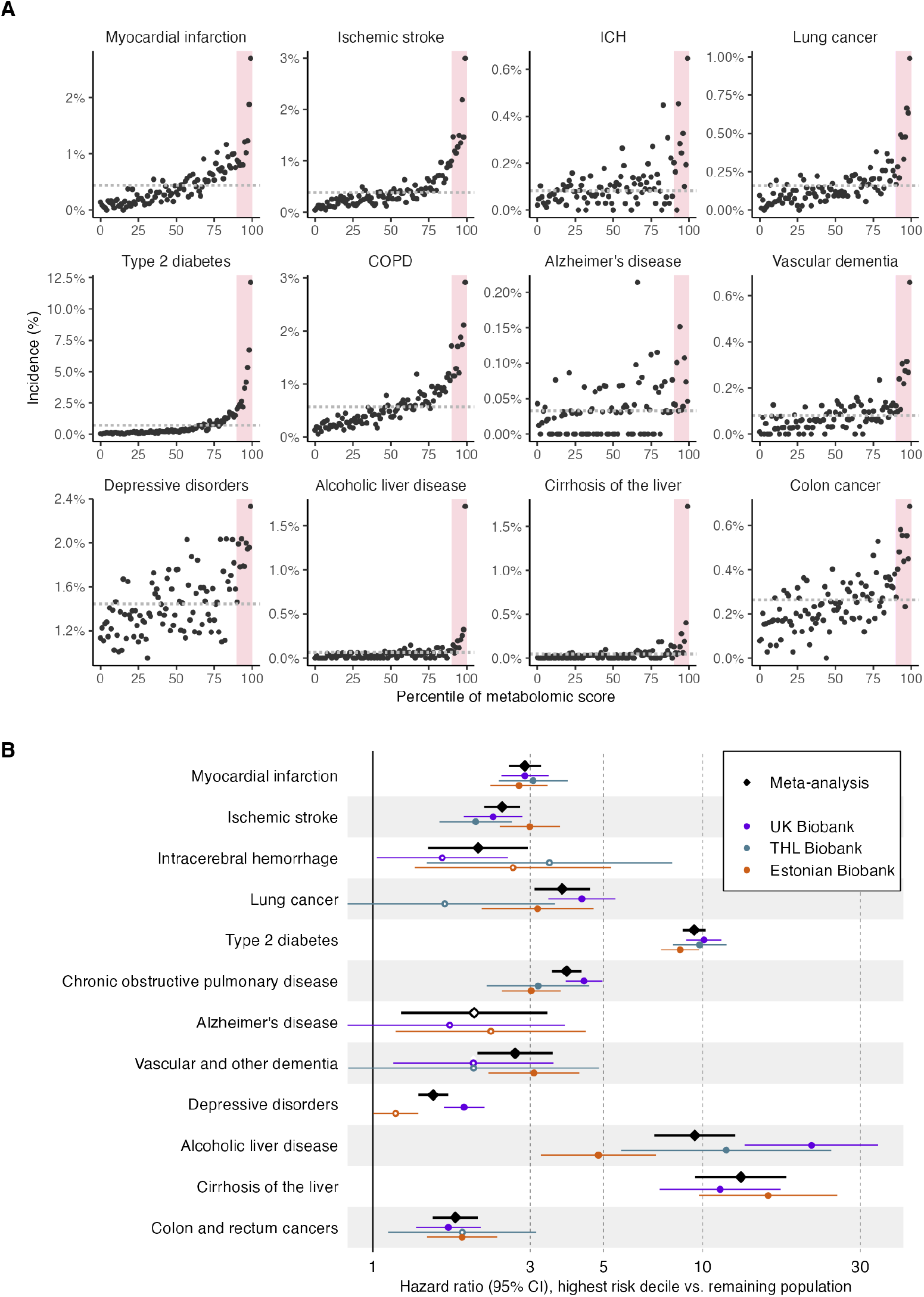
Performance of metabolomic risk scores for future disease onset in three national biobanks. **(A)** Observed incidence of the 12 diseases stratified into one percent bins of the metabolomic score. The observed incidence is shown as a sample size weighted mean of the 4-year incidence in the three biobank cohorts (n=349,418). Red shading shows the top 10% of the metabolomic score (adjusted for age and sex). Horizontal dashed line shows the population prevalence. Metabolomic scores are adjusted for age and sex. **(B)** Hazard ratios of metabolomic scores comparing the highest risk decile to the remaining 90% of the study population for 12 diseases. Horizontal error bars denote 95% confidence intervals.

### Risk model performance

We stratified the three test sets into one percent bins of the risk score distribution and meta-analyzed the four-year incidence rates for each disease (**Fig. 1A**). The risk of disease increased with increasing levels of the metabolomic score across all the diseases studied here. As has been observed previously (*8, 14*), these curves follow a quantile-logistic function, which rises super-exponentially in the tails, making it possible to identify subsets of individuals that are at meaningfully increased risk. This effect is especially dramatic for the scores that most strongly predict disease, including type 2 diabetes and liver disease.

Thus, we hereafter consider the performance of the models using a simplified but plausible preventative public health scenario by comparing the relative four-year risk of incident disease in the 10% of individuals with the highest metabolomic risk scores (“high-risk group”, red shaded area, **Fig. 1A**) to the remaining population. Again, to avoid needing within-cohort comparison data, we used the top decile boundary from our training data to define this group in each of our test sets. The high-risk groups in each biobank had consistently increased risk across diseases (**Fig. 1B**): only depression and alcoholic liver disease showed significant meta-analysis heterogeneity (Cochran’s Q test, p < 0.004 to account for multiple testing). A meta-analysis of the three test sets included hazard ratios of ∼10 for two types of liver disease and diabetes, ∼4 for COPD and lung cancer, and ∼3 for myocardial infarction, vascular dementia and stroke, and was statistically significant (p < 0.004) for all diseases except Alzheimer’s disease (**Table S3**). The pattern of association is similar when considered in standard deviation units, **Fig. S2**.

Furthermore, the UK Biobank test set had the highest point estimate of effect size in only 5 of 12 diseases, demonstrating that the scores are capturing risk factors that are common to multiple countries and eras in time, rather than overfitting to properties specific to the UK Biobank.

Population-wide discrimination, as measured by area under the receiver-operating characteristic curve (AUC), shows consistent, though modest, improvement when adding metabolomic scores on top of age and sex (**Table S3**). While often considered the primary metric of risk model performance, it can be difficult to interpret both absolute AUC and changes between models. Age is a strong risk factor for all these diseases (and a dominating one for some, like dementias), so it is difficult to improve population-wide classification compared to age alone, especially as the three biobanks studied here have widely varying age ranges, and the Estonian Biobank in particular includes many individuals too young to be at meaningful risk of these diseases.

## Combined metabolomic and genetic risk prediction

### Comparative performance of metabolomics and genetics

The observation that a single molecular assay (i.e. metabolomics) can identify individuals at high risk of a wide range of diseases mirrors the concept of polygenic scores (PGS), which have received widespread attention as an opportunity for personalized preventative medicine (*8, 22, 23*). We therefore compared the performance of the metabolomic and polygenic scores. We obtained polygenic scores from the PGS Catalog (**Table S2**) that were built using GWAS data that did not include the UK Biobank, to avoid overfitting. We trained models in half the UK Biobank (now with 10-year follow-up to increase power) always including age and sex, and using Lasso to select from: (i) only the external PGS, (ii) among the metabolomic biomarkers (as above), or (iii) among both the PGS and the metabolomic biomarkers.

PGS were available for 9 of the 12 diseases we considered, and, as expected, the top 10% high-risk groups were at significantly higher risk than the remaining 90% (**Fig. 2A**). However, in all diseases except Alzheimer’s disease and colorectal cancer, the hazard ratio of being in the genetic high-risk group was less than the metabolomic high risk group. In most cases the best performing model included both genetic and metabolomic information, suggesting that these two data types capture at least partially complementary information; a formal interaction test between metabolomic and genetic scores found a significant effect only for type 2 diabetes (**Table S4**). For six diseases we could also calculate PGS in the Estonian Biobank, which replicated the results in the UK Biobank (**Fig S3**).

**Fig. 2.**
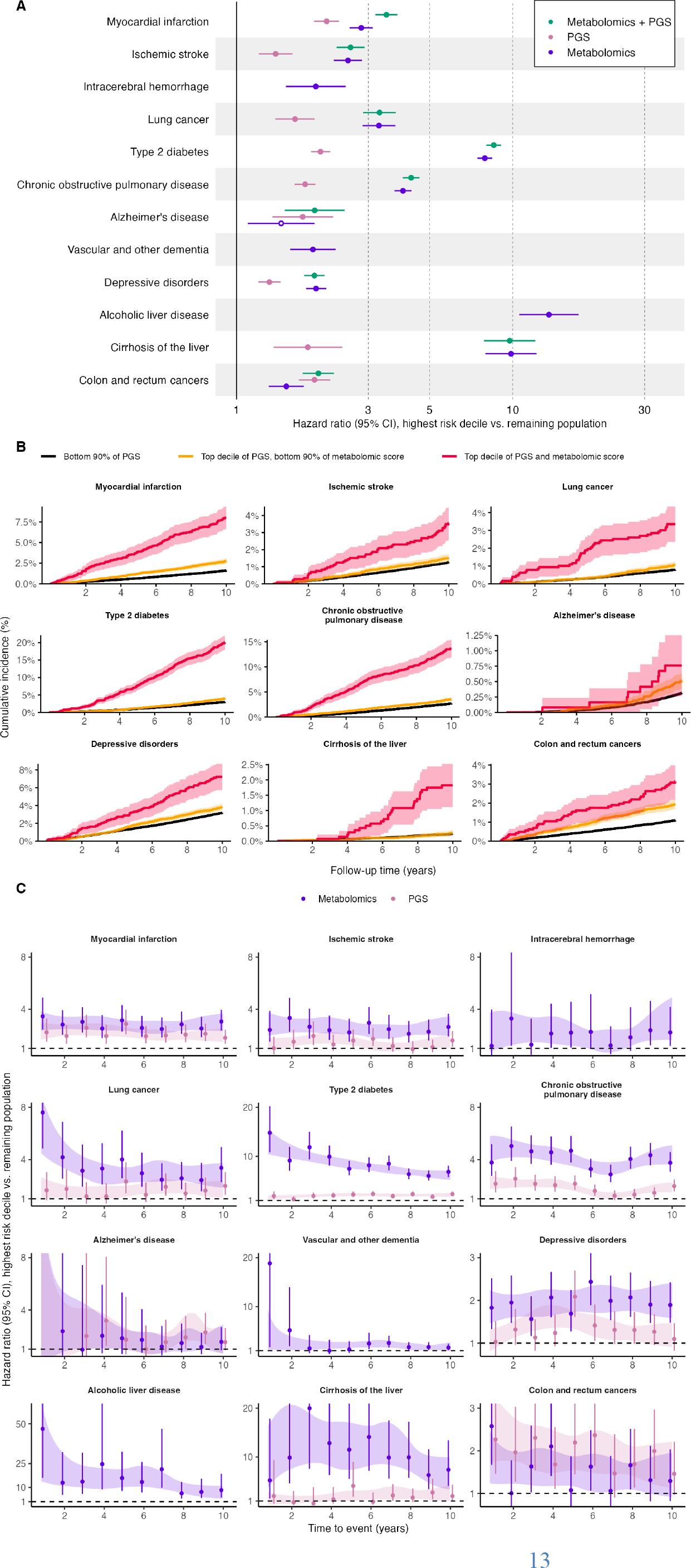
Comparative performance of metabolomic and genetic information on risk. **(A)** Ten-year hazard ratios of metabolomic, polygenic or combined scores comparing the highest risk decile to the remaining study population (UK Biobank test set, n=126,645). Horizontal error bars denote 95% confidence intervals. **(B)** Risk of disease incidence after blood sampling for high genetics risk group stratified by their metabolomic risk score and for average genetic risk group. Shaded region denotes 95% confidence interval. **(C)** Hazard ratios for highest decile of metabolomic or polygenic scores stratified by time to event. Vertical error bars denote 95% confidence intervals per bin, shaded region is 95% confidence interval for a generalized survival model allowing a time-varying effect using natural splines with 2 knots.

We stratified individuals in the genetic high-risk group by whether they were also in the metabolomic high-risk group, or not (**Fig. 2B**). Individuals in both high risk groups are indeed at very elevated risk, but genetically predisposed individuals not in the high metabolomic risk group have risk similar to (or in some cases less than) those not in the genetically predisposed group. This effect is likely due to a combination of factors: current PGS do not actually capture most of the genetic risk for these diseases, and that unexplained heritability, combined with lifestyle and environmental history is partially reflect in the metabolomic score. The metabolomic and genetic scores also have different patterns of correlation between different diseases (**Fig S4**). As has been previously shown, the PGS tend to be largely uncorrelated (*23*), whereas the metabolomic scores are nearly all correlated with each other, reflecting how they capture “multi-morbidity” (*14, 17*). Combining the two types of information can yield both improved discrimination and greater specificity of risk stratification.

While the three biobanks are dominated by individuals of European ancestry, we did compare the transferability of the metabolomic scores and PGS for five endpoints with at least 35 events in multiple ancestries in the UK Biobank (**Fig. S5)**. The metabolomic scores remained significantly predictive across disease-ancestry combinations, though often with weaker effect size estimates than in the European ancestry group. As has been previously shown, the effect sizes of PGS were also attenuated in non-European ancestries, and because they do not predict as well to begin with in Europeans, the estimate was not statistically significant in 11 of 14 non-European comparisons. This suggests that metabolomic scores may presently have more value in non-European ancestries than PGS, but additional, more diverse datasets will be essential to produce risk scores that are as widely useful as possible.

The longer 10 years of follow-up in the UK Biobank also allowed us to compare short-term and long-term prediction from these scores. As expected, since PGS risk is fixed throughout life, PGS hazard ratios remained constant over time (**Fig. 2C**). The relationship between metabolomic risk and time to event varied by disease: the type 2 diabetes, lung cancer and alcoholic liver disease scores provide stronger stratification of near-term risk. For most diseases, however, the metabolomic risk scores were stable over time, similar to PGS. In all cases the better overall score was consistently better across 10 years.

## Metabolomic prediction from multiple timepoints

We generated metabolomic profiles at a second time point from blood samples donated by 13,817 UK Biobank participants who returned for a repeat visit approximately five years after they initially enrolled in the study. The correlations of the scores range from 0.45 for Alzheimer’s disease to 0.72 for myocardial infarction and diabetes, which fall in the middle of the range of correlations for individual biomarkers (e.g. amino acids ∼ 0.2, HDL cholesterol ∼ 0.8) (**Table S5, Fig. S6**). This level of stability reinforces the observation above that the scores provide information about disease risk 10 years into the future, but since the correlation is not perfect it also implies that multiple measurements will provide more complete information for risk prediction.

For six diseases (myocardial infarction, ischemic stroke, diabetes, COPD, depression, and colorectal cancer) at least 100 events occurred within 10 years of the repeat visit, so we fitted a joint risk model with baseline and follow-up metabolomic score measurements. For myocardial infarction (p_baseline_ = 6.7×10^−3^, p_follow-up_ = 6.7×10^−7^), diabetes (p_b_ = 3.4×10^−12^, p_f_ = 6.8×10^−34^) and COPD (p_b_ = 5.7×10^−6^, p_f_ = 8.3×10^−10^) both time points were significantly associated with 10-year risk; for the other three diseases the hazard ratio point estimates were all consistently positive but were not significant due to weaker prediction from the scores and smaller sample size. This suggests that our scores are not forgetful: both a person’s currently assessed metabolomic risk, and information about how long they have lived in a state of elevated risk, contribute information about risk of future disease onset after the second time point.

To further explore this idea, we considered individuals in the top 10% high risk groups for myocardial infarction, diabetes, or COPD at the first time point, and compared the subset of that group who remained in the high-risk group at the follow-up time point to those who had left it. For diabetes and COPD, changing risk strata showed a dramatic reduction in future risk (3.1-fold and 2.4-fold, p=3×10^−6^ and 0.002, respectively), after adjusting for baseline risk (**Fig. 3**). We replicated this analysis in 5,202 individuals from the Estonian Biobank for whom we had similarly profiled a second sample from approximately five years after the first. We observed the same effect for type 2 diabetes, which was the only disease for which we had sufficient cases to test it (HR=4.5, p=0.002).

**Figure 3.**
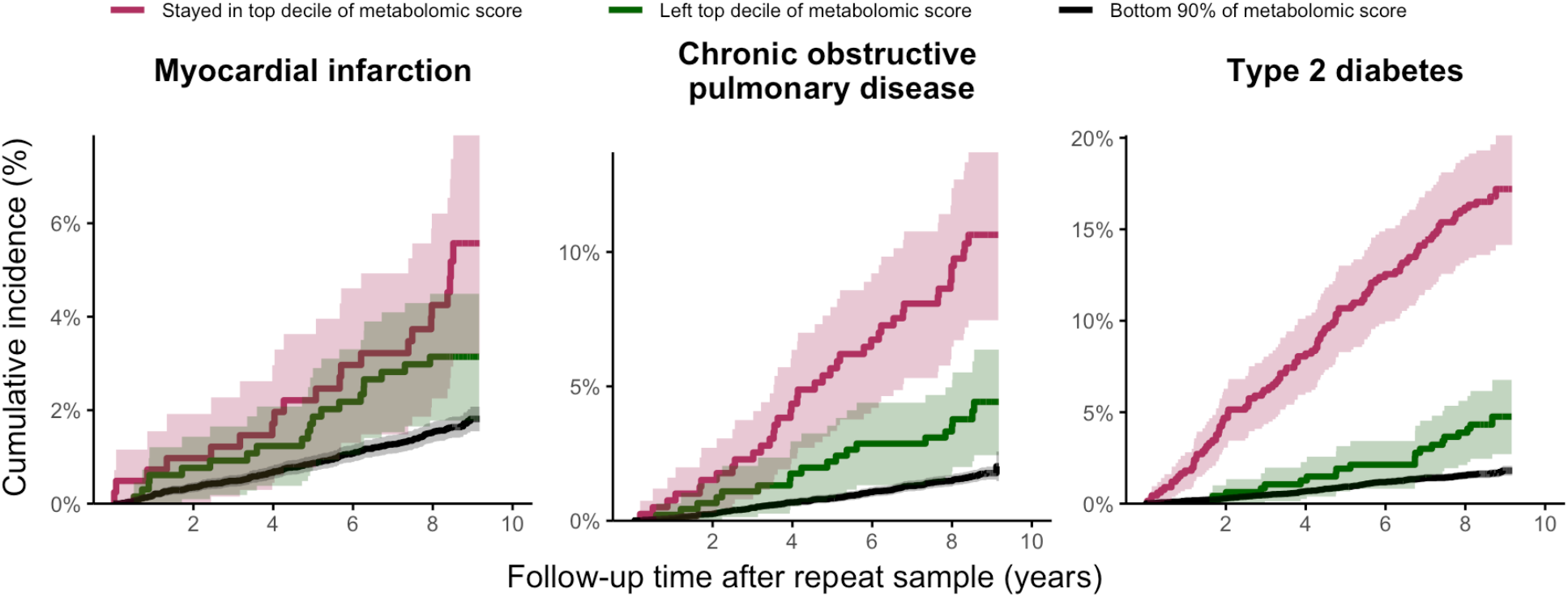
Risk of future disease onset for three diseases among those who remain in the high-risk group over multiple years, or leave it. Maroon lines show those who were in the high-risk group at both time points ∼5 years apart, green lines show those who were in the high-risk group at enrolment but had left it by the second time point, black lines show those who were not in the high-risk group at enrolment.

While we do not know what caused individual metabolomic scores to change between time points, we can assess what differences in lifestyle factors are associated with changes in metabolomic risk scores. For example, obese individuals who stayed in the high-risk group for diabetes gained an average 0.21 units of BMI, but those who switched from high to low risk, lost an average of 0.60 units of BMI (difference of 0.81, p = 3×10^−14^). Of self-reported smokers who were in the high-risk COPD group at the first time point, 65% of those who continued smoking remained at high risk, compared to just 40% of those who reported quitting between the two time points (p=0.004). There were also associations to several other lifestyle variables, including amount of exercise, alcohol consumption, and dietary habits. However, these collectively explained only a few percent of the observed metabolomic score changes, demonstrating that the scores integrate a much wider range of information than questionnaires.

## Real-world multi-omic prediction

### Clinical characteristics of individuals with high metabolomic risk

We next sought to understand how ‘omic prediction scores relate to the kinds of information that are likely available in existing preventative healthcare settings, such as visits to primary care or national health check programs. First, we compared our high-risk groups to the remainder of the population using the Frailty Index (*24, 25*) as a surrogate for a primary care physician’s overall impression of the health of an individual. As expected, the high-risk group have slightly higher frailty index values (**Fig. S7, Table S6**). However, the difference cannot routinely identify individuals in the high-risk groups. Second, we considered the relative performance of a simple risk model using commonly available clinical data in preventative health: blood pressure, total and HDL cholesterol, BMI, and smoking status. The ‘omic predictions indeed outperform these, and a model where the cholesterol test component of the standard clinical data is replaced with ‘omic data performs dramatically better (**Fig. 4A**). Lung cancer is a special case, since it is so closely tied to smoking status (and being a smoker essentially determines membership in the high-risk group). When considering the performance of the scores only in smokers, the omics score has a hazard ratio of 2.8, compared to 1.2 for the clinical score.

**Fig. 4.**
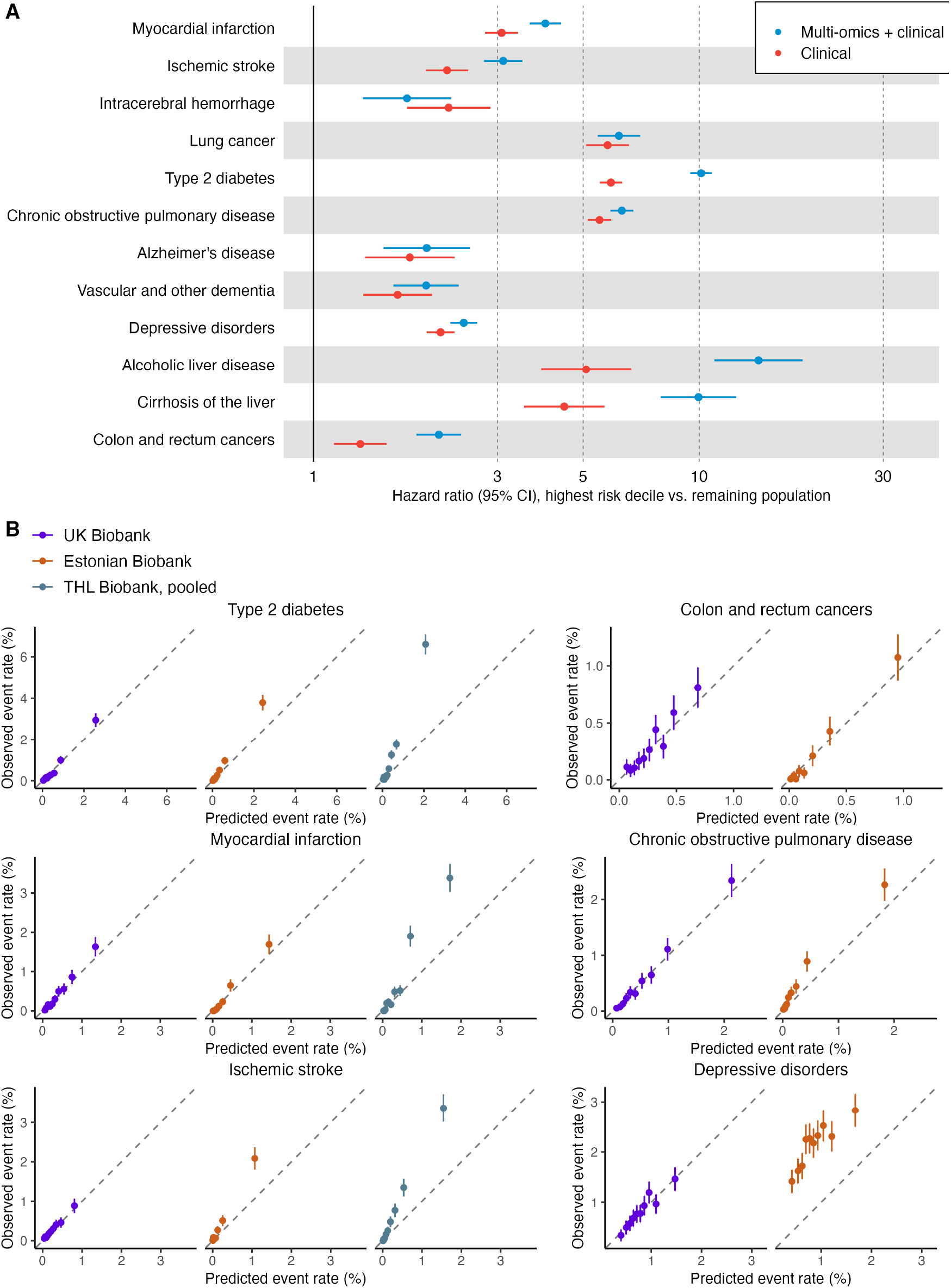
Implementing omic prediction in public health settings. **(A)** Performance of a simple model with clinical variables compared to those variables plus the best ‘omic data (either just metabolomics or metabolomics plus PGS from Figure 2). **(B)** For each disease, the calibration of three-year observed event rates are shown by 10 equally sized deciles of absolute predicted risk. Vertical lines represent 95% confidence intervals. Calibration slopes and intercepts were derived from a logistic regression of the observed risk on the predicted risk.

### Calibration across studies from different countries

Finally, real-world use of any risk prediction model requires both discrimination and calibration (*26*). We therefore tested the calibration of our metabolomic scores by plotting observed event rates against predicted absolute event rates per decile in all three biobanks (**Fig. 4B**). We estimated calibration slopes and intercepts by fitting a logistic regression model of observed risk on predicted risk, without any study-specific processing or normalization, mimicking real-world usage (**Methods**). For the main calibration analysis, we included diseases with >200 events over 3 years, as recommended by earlier studies (*27, 28*). Calibration results for the remaining diseases are shown in **Figure S8**.

Overall, the metabolomic risk scores demonstrated good calibration. In UK Biobank test set the calibration slopes ranged from 0.94 to 1.17 across diseases, as expected since the models were trained in the other half of this biobank. In the independent Estonian Biobank, the calibration slopes ranged from 0.78 to 1.15, except for depression at 0.47. This difference is likely a result of diagnostic differences in depression in different countries, as well as how those diagnoses are encoded in electronic records. In THL Biobank the slopes were 1.03 (ischemic stroke), 1.20 (myocardial infarction) and 1.21 (diabetes), possibly reflecting different rates of these diseases in the earlier recruitment waves of these cohorts.

## Discussion

Here we have shown that metabolomic risk scores can identify a small group of individuals at meaningfully increased risk (here we considered the highest risk decile, but different cut points will be appropriate for different diseases). Indeed, focusing on a defined high-risk group, without considering the relative stratification across the entire population (e.g. via AUC) is how many existing preventative health initiatives are designed: if an individual’s absolute risk is above a certain threshold, a pre-defined action is recommended. We have shown that the high-risk groups have consistently increased risks in biobanks from three countries which have varying sample types (plasma vs. serum), study designs, and fasting protocols. The calibration results, while imperfect, are comparable to widely used tools like the pooled cohort equations for cardiovascular risk when compared, for example between the US and Canada (*29*).

We believe that personalized prevention using genetics, which has generated substantial excitement as a sea-change in medicine, should be paired wherever possible with metabolomic risk scores, as the prediction is better than currently available PGS for 10 of 12 diseases examined here. When good predictors from both data types exist, such as for myocardial infarction and diabetes, the data are complementary, and together provide an optimum combination of predictive accuracy and specificity for individuals interested in reducing their risk of specific diseases, rather than just multi-morbidity, which may seem less actionable.

Communicating polygenic risk to both healthcare practitioners and individuals is also challenging (*30*), and individuals presented with information that they have high polygenic risk for a disease may conclude their “bad genes” mean they are destined to develop the disease. Metabolomics offers a two-fold solution: first by clearly showing that an individual’s lifestyle to date strongly affects their risk of disease, enabling a positive message, and second, by studying >18,000 individuals in this dataset with two metabolomic measurements five years apart we can show that the scores have an attractive balance of stability and responsiveness to lifestyle.

Can these approaches be incorporated into public health practice? While some new technologies, like proteomics (*31*), provide even more powerful risk stratification, their implementation faces challenges because they do not yet provide absolute quantifications. Metabolomic risk scores using NMR data, by contrast, can be paired with familiar individual biomarker levels (e.g. cholesterol and glucose). Metabolomic scores are stable for multiple years of follow-up, and can readily fit into 2- or 5-year public health check plans, especially when generated using blood samples that are already part of routine clinical sample flows (e.g. by replacing a cholesterol test with a full metabolomic measurement at only marginally increased cost). And, considering cardiovascular diseases, lung diseases, liver diseases and diabetes, where the high-risk group has at least 3-fold increase in risk, and where interventions already exist, 28% of individuals in the UK Biobank are in the high-risk group for at least one disease (likely an underestimate of population levels, due to healthy volunteer bias). This combination of achievable implementation and widespread potential impact suggests that it is now possible to use multi-omic prediction in clinical practice.

## Data Availability

The Nightingale Health NMR biomarker data for 300,000 individuals are released to the UK Biobank resource in July 2023 (https://biobank.ndph.ox.ac.uk/showcase/label.cgi?id=220). The UK Biobank data are available for approved researchers through the UK Biobank data-access protocol (https://www.ukbiobank.ac.uk/enable-your-research/apply-for-access). Data from Estonia Biobank can be accessed through a research application to Institute of Genomics of the University of Tartu (https://genomics.ut.ee/en/content/estonian-biobank). Data from FINRISK and Health 2000 cohorts can be accessed through a research application to THL Biobank (https://thl.fi/en/web/thl-biobank).

https://www.ukbiobank.ac.uk/enable-your-research/apply-for-access

https://genomics.ut.ee/en/content/estonian-biobank

https://thl.fi/en/web/thl-biobank

## Acknowledgments

We acknowledge the lab and spectrometry teams at Nightingale Health for their role in generating the metabolomic data on these cohorts. We are grateful to UK Biobank (Project 30418), Estonian Biobank, and THL Biobank (project BB2016_86) for access to data to undertake this study. We thank all biobank participants for their generous contribution to generating this resource for the scientific community. The Estonian Biobank Research Team consists of Mari Nelis, Georgi Hudjasov, Reedik Mägi, Andres Metspalu, and Lili Milani.

## Author contributions

Conceptualization: JCB, TE, HJ, PW

Data Curation: HJ, NK, SK, SNL, KS

Formal Analysis: LJ-D, HJ, NK, SK, SNL, KS

Methodology: HJ

Investigation: JCB, LJ-D, HJ, KH, NK, SK, AK, SNL, VM, KN, KS, MS, PS, MT, PW

Resources: TE, PJ, TJ, JK, AL, MP, VS, TT

Supervision: JCB, HJ, PW

Visualization: LJ-D, HJ, SK, SNL, KS

Writing – original draft: JCB

Writing – review & editing: JCB, LJ-D, HJ, SK, SNL, NK, KS, PW

## Supplementary Materials

Materials and Methods

Figs. S1 to S8

Tables S1 to S5

