## Supplementary Materials for "Metabolomic and genomic prediction of common diseases in 477,706 participants in three national biobanks"

Nightingale Health Biobank Collaborative Group

**The PDF file includes:**

Materials and Methods
Figs. S1 to S8
Tables S1, S4, S5, S6
References

**Other Supplementary Materials for this manuscript include the following:**

Tables S2 to S3

### Materials and Methods

Study populations and endpoint definitions

We used data from a total of 477,706 individuals from three biobanks, UK Biobank (N = 254,993), Estonian Biobank (N = 190,785) and Finnish THL Biobank (N = 31,928). Written informed consent was obtained from all participants.

The UK Biobank is a longitudinal biomedical study of approximately half a million participants between 38-71 years old from the United Kingdom (*1*). Participant recruitment was conducted on a volunteer basis and took place between 2006 and 2010. Initial data were collected in 22 different assessment centers throughout Scotland, England, and Wales. Data collection includes elaborate genotype, environmental and lifestyle data. Blood samples were drawn at baseline for all participants, with an average of four hours since the last meal, i.e. generally non-fasting. NMR metabolomic biomarkers (Nightingale Health, quantification library 2020) were measured from EDTA plasma samples (aliquot 3) during 2019–2022 from a random subset of ~275,000 individuals out of the entire cohort. In addition, plasma samples were measured by NMR metabolomics from approximately 15,000 participants (out of 20,000 enrolled) who underwent a repeat-visit assessment on average five years after the baseline visit. Details on the NMR metabolomic measurements in UK Biobank have been described previously for the first tranche of ~120,000 samples (*2*). This study represents the release of the second tranche of NMR metabolomic data in the UK Biobank, released for the research community via UK Biobank access procedures in July 2023. The NMR metabolomic measurements from the entire baseline cohort are underway. Follow-up data include a wide range of electronic health-related records, including disease incidence, hospital admissions, primary care, and death records, which are presently still regularly updated. The UK Biobank study was approved by the North West Multi-Centre Research Ethics Committee. This research was conducted using the UK Biobank Resource under Application Number 30418.

The Estonian Biobank (EBB) is a curated population-based biobank of Estonia, comprising a cohort of approximately 210,000 individuals (*3*). The enrollment was primarily based on volunteering rather than ensuring representativeness. Nonetheless, the participants constitute about 20% of the adult Estonian population and the cohort is representative of the nation in terms of age, sex, and geographic dispersion. The enrollment was conducted between 2002 and 2022. A network comprising general practitioners and various medical personnel from private practices, hospitals, and recruitment offices of the Estonian Genome Center was established for participant recruitment, as well as for collection of samples and health data. After recruitment, participants were asked to fill out detailed questionnaires, which encompassed personal information, genealogical data, educational and occupational history, as well as lifestyle habits. Blood samples were generally collected non-fasting (*4*). NMR metabolomic measurements were conducted on EDTA plasma samples (100 uL) for all biobank participants. The EBB database undergoes regular synchronization with several national registries and hospital databases, along with the national health insurance fund's database that houses comprehensive treatment and service bill information. Disease events are codified in compliance with the ICD-10 standards and medication usage is categorized as per the Anatomical Therapeutic Chemical (ATC) classification, both with current follow up data available until the end of 2021. The Estonian Committee on Bioethics and Human Research approved the study. Data was accessed with research approval number 1.1-12/2770.

The Finnish THL biobank data consist of five population cohorts (National FINRISK Studies 1997, 2002, 2007, 2012 and Health 2000 Survey) collected in study specific years between 1997 and 2012 (*5*, *6*). Each of the five cohorts is an independent random sample drawn of unique individuals aged 25-98 (25–74 in FINRISK, 30 and over in Health 2000). Recruitment was conducted via invitation-only in multiple urban and rural areas across Finland to be representative of the nation (participant rate 60-70%). The baseline surveys included a wide range of health-related questionnaire and biological measures, including a non-fasting blood sample (median 5 hours since last meal) for biomarker measurements from ~85% of all participants enrolled. NMR metabolomic data were measured from all participants with blood samples available. In contrast to UK and Estonian Biobanks, NMR metabolomics measurements were done on serum samples (350 uL), as described in detail previously (*2*, *5*). Information on disease outcomes were linked from national hospital discharge registries and reimbursement records with follow-up until 2017 (4 to 19 years of follow-up). The THL biobank cohorts were approved by the Coordinating Ethical Committee of the Helsinki and Uusimaa Hospital District, Finland. Data was accessed with research application number BB2016_86.

In our main analysis, we included 12 diseases that are among the top causes of disease adjusted life years (DALYs) in the European region according to the WHO (*7*): myocardial infarction, ischemic stroke, intracerebral hemorrhage, lung cancer, type 2 diabetes, chronic obstructive pulmonary disease (COPD), Alzheimer disease, vascular and other dementias, depressive disorders, alcoholic liver disease, cirrhosis of the liver, and colon and rectum cancers. We split up or specified some of the original WHO groupings to allow for more focused risk prediction and be aligned with common disease definitions used for PGS development. Instead of using ischemic heart disease and diabetes mellitus, we specified to myocardial infarction and type 2 diabetes, stroke was split into ischemic stroke and intracerebral hemorrhage, Alzheimer's disease was separated from vascular and other dementias, and alcoholic liver disease was separated from cirrhosis of the liver.

Disease incidence was defined based on the first occurrence of ICD-10 codes listed in **Table S2**. In the UK Biobank, first occurrences were based on primary care data (only available in approximately 45% of UK Biobank participants), hospital inpatient data, death register records, cancer registry data, and self-reports at baseline. In the Estonian Biobank, first occurrences were based on self-reports at baseline, E-Health, North Estonia Medical Center, Tartu University Hospital, death registry records and cancer registry data. The Estonian Health Insurance Fund was excluded as a source. We considered only incident cases occurring after the blood draw at study baseline. Prevalent cases were excluded from the analysis in a disease-specific manner.

Metabolomic biomarker profiling

Lipid and metabolite biomarkers were quantified from approximately 500,000 blood samples by high-throughput NMR metabolomics (Nightingale Health Ltd). Details of the sample handling and measurement protocol for UK Biobank have been described in detail (*2*). The measurement protocol was similar for Estonian Biobank and Finnish THL Biobank (*5*). Briefly, the samples were profiled using a total of nine 500 MHz spectrometers (Bruker AVANCE IIIHD). Two NMR spectra are recorded and proprietary software is used for biomarker quantification in absolute units (Nightingale Health, quantification library 2020). (*2*, *8*). This provides 249 biomarker measures in a single assay (168 absolute and 81 ratio measures), including routine lipids, lipoprotein profiling of 14 size subclasses, fatty acids, and various low-molecular weight metabolites, such as amino acids, ketones, and glycolysis metabolites as well as two inflammatory protein measures, albumin, and glycoprotein acetyls. These metabolomic biomarker data from ~300,000 samples are now available to approved researchers through the UK Biobank for all public health research. To obtain the most robust possible risk models, we used 36 clinically validated biomarkers with diagnostics approval in the NMR metabolomics assay to facilitate rapid translation and clinical applications for model training (Total cholesterol, VLDL cholesterol, Clinical LDL cholesterol, HDL cholesterol, Total triglycerides, Apolipoprotein B, Apolipoprotein A1, Ratio of apolipoprotein B to apolipoprotein A1, Total fatty acids, Omega-3 fatty acids, Omega-6 fatty acids, Polyunsaturated fatty acids, Monounsaturated fatty acids, Saturated fatty acids, Docosahexaenoic acid, Ratio of omega-3 fatty acids to total fatty acids, Ratio of omega-6 fatty acids to total fatty acids, Ratio of polyunsaturated fatty acids to total fatty acids, Ratio of monounsaturated fatty acids to total fatty acids, Ratio of saturated fatty acids to total fatty acids, Ratio of docosahexaenoic acid to total fatty acids, Ratio of polyunsaturated fatty acids to monounsaturated fatty acids, Ratio of omega-6 fatty acids to omega-3 fatty acids, Alanine, Glycine, Histidine, Total concentration of branched-chain amino acids, Isoleucine, Leucine, Valine, Phenylalanine, Tyrosine, Glucose, Creatinine, Albumin, Glycoprotein acetyls) (*2*) . To account for potential glucose degradation prior to plasma sample preparation, we used an estimate of physiological glucose concentration based on observed glucose and lactate as input for the risk models. Further, to correct for spectrometer differences in alanine concentration, the single metabolite most impacted by technical variation (*9*), we shifted mean alanine concentrations observed within each spectrometer in each biobank to the mean and standard deviation of a master spectrometer. The average biomarker detection rate was >99% across the plasma samples. Further details on the individual biomarker measures are provided in the UK Biobank data resource.

Genotype data and polygenic scores

For this study, genotype data was available for UK Biobank and Estonian Biobank, but not for THL Biobank. The UK Biobank participant have been genotyped on Applied Biosystems UK Biobank Axiom Array and UK BiLEVE Axiom Array, measuring over 800,000 variants and imputed using the Haplotype Reference Consortium and UK10K and 1000 Genomes reference panels outside of this study (*1*). Estonian Biobank participants have been genotyped with genome-wide chip arrays and further imputed with a population-specific imputation panel consisting of high-coverage (30-fold) whole-genome sequence data from 2,244 individuals and over 16 million high-quality genetic variants (*10*).

For 9 of the 12 diseases, we used an existing polygenic score (PGS) that was developed using genome-wide association study (GWAS) summary statistics that did not include the UK Biobank in their discovery cohort (**Table S2**). These PGS were computed for UK Biobank and Estonian Biobank participants as the weighted sum of risk alleles using imputed genotype data. We acknowledge that a small number (~8,000) of Estonian Biobank samples may have been included in the GWAS underlying the PGS of diabetes and myocardial infarction. For UK Biobank, we estimated participant's genetic ancestry with respect to the five superpopulations of the 1000 Genomes Project (*11*) using principal component analysis projection and a random forest classifier, and scaled PGS with respect to their estimated ancestry. For Estonian Biobank we assumed more homogenous genetic background and scaled PGS within the cohort.

Statistical analyses

Prior to epidemiological data analyses, we initially excluded samples with any missing metabolomic biomarkers (8284 samples in UK Biobank; 3.0%) and subsequently samples with at least one concentration value more than the mean ± 4 standard deviations (9284 samples in UK Biobank; 3.5%). Additionally, we excluded samples lacking basic risk factor information or genetic data. We did not filter on genetic ancestry or ethnicity. Metabolomic biomarker measures were log1p-transformed, and all continuous variables were Z-normalized to have a mean of zero and a standard deviation of one in the training set. The means and standard deviations of the training set were subsequently used to scale the metabolomic biomarkers in the validation and replication cohorts.

We trained risk models using a randomized 50% of the UK Biobank data, excluding participants with a repeat measurement (N=120,123), with 10-years of follow-up and Cox proportional hazards regression modeling with least absolute shrinkage and selection operator (LASSO) and five-fold cross-validation. For each disease, we trained four models, which included (1) age and sex, (2) age, sex, and metabolomic measures, (3) age, sex, and a disease-specific PGS, and (4) age, sex, metabolomic measures, and a disease-specific PGS. Age and sex were not penalized to ensure they were selected for each model and appropriately weighted. For the genetic ancestry analyses, we trained another set of models identical to the description above but leaving all participants with non-European genetic ancestry out from the training set to maximize the size of this group in the testing set. We additionally trained a set of three clinical models that included (1) age, sex, smoking status, BMI, systolic blood pressure, HDL cholesterol, and total cholesterol (basic clinical variables), (2) basic clinical variables and metabolomic data, and (3) basic clinical variables, metabolomic data, and a disease-specific PGS.

Risk scores were computed as the weighted sum of selected predictors for each model. In computing the risk scores, we excluded the sex and age coefficients, which for a combined metabolomic and PGS model for example, effectively results in risk scores comprised of the weighted sum of 1-36 metabolomic measures and a PGS, which have been adjusted for age and sex.

Risk model performance was tested by assessing the association of these age- and sex-adjusted scores with disease incidence within the first 4 years after the blood draw using Cox proportional hazards models and examining the hazard ratios (HR) with 95% confidence intervals between individuals in the top decile of the score and individuals at the bottom 90%. The limits for the top deciles for each model were calculated in the training set once and subsequently used to determine top decile classification in validation and replication cohorts. We similarly assessed HR for 10 years of follow-up. In addition, we computed HR per one standard deviation increments in the age- and sex-adjusted scores using Cox proportional hazards models. Correlations of the scores were evaluated by computing Pearson correlation coefficients. For statistical significance, we considered p-values $< 0.004$ corresponding to a 95% confidence level Bonferroni corrected for 12 diseases (*12*, *13*). Kaplan-Meier curves stratified by PGS and metabolomic score deciles were estimated for 10 years of follow-up using R package survival (*14*) for individuals in the top decile of the PGS score and the bottom 90% of the metabolomic score, the top decile of both the PGS and metabolomic scores, and the bottom 90% of the PGS score. We used the cox.zph function to examine the proportionality of hazards assumption and Schoenfeld residual plots, which revealed that the HR was not constant over the 10-year follow-up time. Therefore, we assessed the continuous hazard over the HR in strata across the follow-up time for the metabolomics and PGS models. Using the R package rstpm2, we built a generalized survival model with natural splines using 2 knots to allow for a time-varying effect, and additionally computed the hazard ratio for 1-year strata using the survSplit option.

For analyses examining two time points, we started with 14,939 UKB participants with metabolomic measures both from baseline visit and a repeat visit which took place after 2 to 7 years. Of these, we excluded 1,122 participants who had either incomplete or outlying (over 4 standard deviations from the mean) metabolite measures. For the following analyses, we considered the six diseases that had at least 100 cases within 10 years after the repeat visit among the 13,817 individuals left for the analyses (273 cases for COPD, 164 for colon cancer, 231 for depression, 331 for diabetes, 247 for myocardial infarction and 179 ischemic stroke). To explore the effects of the two different time points, we fitted Cox proportional hazards models explaining diseases events 10 years after the repeat visit with age- and sex-adjusted metabolic scores at baseline and repeat visit. New disease events between baseline and repeat visit were excluded. To assesses the risk changes between the visits, we categorized participants into three groups: those who stayed in the highest decile of metabolic risk, those who left the highest decile, and the remaining population and compared their risk. Additionally, we analyzed 5,202 participants with two separate blood measurement time points from the Estonian Biobank as above.

We assessed clinical characteristics of high-risk individuals, defined as those with a metabolomics model score in the highest decile of at least one of the seven best-performing models: alcoholic liver disease, chronic obstructive pulmonary disease, cirrhosis of the liver, ischemic stroke, lung cancer, myocardial infarction, and type 2 diabetes. In addition to basic clinical characteristics, we evaluated the frailty index, a measure to quantify aging and health, between high- and low-risk individuals. We calculated the frailty index based on the method described by Williams et al. (*7*), using 49 self-reported disease outcomes in UK Biobank participants. Participants with at least ten missing items were excluded. We tested the difference in mean for clinical characteristics between high- and low-risk individuals using a two-sided t-test for continuous variables and a Chi-squared test for categorical variables.

To evaluate calibration of the metabolomic scores, we estimated observed and predicted incidence rates in all three biobanks over 3 years of follow-up. Censoring was chosen to be at three years to obtain complete and comparable follow-up for as many samples as possible. Calibration slopes and intercepts were estimated by fitting logistic regression of individual diseases status (observed risk) on predicted risk (*15*, *16*).

All statistical analyses and modeling were performed in R version 4.2.

4. K. Fischer, J. Kettunen, P. Würtz, T. Haller, A. S. Havulinna, A. J. Kangas, P. Soininen, T. Esko, M.-L. Tammesoo, R. Mägi, S. Smit, A. Palotie, S. Ripatti, V. Salomaa, M. Ala-Korpela, M. Perola, A. Metspalu, Biomarker profiling by nuclear magnetic resonance spectroscopy for the prediction of all-cause mortality: an observational study of 17,345 persons. *PLoS Med*. **11**, e1001606 (2014).
5. E. Tikkanen, V. Jägerroos, M. V. Holmes, N. Sattar, M. Ala-Korpela, P. Jousilahti, A. Lundqvist, M. Perola, V. Salomaa, P. Würtz, Metabolic Biomarker Discovery for Risk of Peripheral Artery Disease Compared With Coronary Artery Disease: Lipoprotein and Metabolite Profiling of 31 657 Individuals From 5 Prospective Cohorts. *J Am Heart Assoc*. **10**, e021995 (2021).
6. K. Borodulin, H. Tolonen, P. Jousilahti, A. Jula, A. Juolevi, S. Koskinen, K. Kuulasmaa, T. Laatikainen, S. Männistö, M. Peltonen, M. Perola, P. Puska, V. Salomaa, J. Sundvall, S. M. Virtanen, E. Vartiainen, Cohort Profile: The National FINRISK Study. *International Journal of Epidemiology*. **47**, 696–696i (2018).
7. World Health Organization, Global health estimates: Leading causes of DALYs, (available at https://www.who.int/data/gho/data/themes/mortality-and-global-health-estimates/global-health-estimates-leading-causes-of-dalys).
8. P. Soininen, A. J. Kangas, P. Würtz, T. Suna, M. Ala-Korpela, Quantitative Serum Nuclear Magnetic Resonance Metabolomics in Cardiovascular Epidemiology and Genetics. *Circulation: Cardiovascular Genetics*. **8**, 192–206 (2015).
9. S. C. Ritchie, P. Surendran, S. Karthikeyan, S. A. Lambert, T. Bolton, L. Pennells, J. Danesh, E. Di Angelantonio, A. S. Butterworth, M. Inouye, Quality control and removal of technical variation of NMR metabolic biomarker data in ~120,000 UK Biobank participants. *Sci Data*. **10**, 64 (2023).
10. M. Mitt, M. Kals, K. Pärn, S. B. Gabriel, E. S. Lander, A. Palotie, S. Ripatti, A. P. Morris, A. Metspalu, T. Esko, R. Mägi, P. Palta, Improved imputation accuracy of rare and low-frequency variants using population-specific high-coverage WGS-based imputation reference panel. *Eur J Hum Genet*. **25**, 869–876 (2017).
11. A. Auton, G. R. Abecasis, D. M. Altshuler, R. M. Durbin, G. R. Abecasis, D. R. Bentley, A. Chakravarti, A. G. Clark, P. Donnelly, E. E. Eichler, P. Flicek, S. B. Gabriel, R. A. Gibbs, E. D. Green, M. E. Hurles, B. M. Knoppers, J. O. Korbel, E. S. Lander, C. Lee, H. Lehrach, E. R. Mardis, G. T. Marth, G. A. McVean, D. A. Nickerson, J. P. Schmidt, S. T. Sherry, J. Wang, R. K. Wilson, R. A. Gibbs, E. Boerwinkle, H. Doddapaneni, Y. Han, V. Korchina, C. Kovar, S. Lee, D. Muzny, J. G. Reid, Y. Zhu, J. Wang, Y. Chang, Q. Feng, X. Fang, X. Guo, M. Jian, H. Jiang, X. Jin, T. Lan, G. Li, J. Li, Y. Li, S. Liu, X. Liu, Y. Lu, X. Ma, M. Tang, B. Wang, G. Wang, H. Wu, R. Wu, X. Xu, Y. Yin, D. Zhang, W. Zhang, J. Zhao, M. Zhao, X. Zheng, E. S. Lander, D. M. Altshuler, S. B. Gabriel, N. Gupta, N. Gharani, L. H. Toji, N. P. Gerry, A. M. Resch, P. Flicek, J. Barker, L. Clarke, L. Gil, S. E. Hunt, G. Kelman, E. Kulesha, R. Leinonen, W. M. McLaren, R. Radhakrishnan, A. Roa, D. Smirnov, R. E. Smith, I. Streeter, A. Thormann, I. Toneva, B. Vaughan, X. Zheng-Bradley, D. R. Bentley, R. Grocock, S. Humphray, T. James, Z. Kingsbury, H. Lehrach, R. Sudbrak, M. W. Albrecht, V. S. Amstislavskiy, T. A. Borodina, M. Lienhard, F. Mertes, M. Sultan, B. Timmermann, M.-L. Yaspo, E. R. Mardis, R. K. Wilson, L. Fulton, R. Fulton, S. T. Sherry, V. Ananiev, Z. Belaia, D. Beloslyudtsev, N. Bouk, C. Chen, D. Church, R. Cohen, C. Cook, J. Garner, T. Hefferon, M. Kimelman, C. Liu, J. Lopez, P. Meric, C. O'Sullivan, Y. Ostapchuk, L. Phan, S. Ponomarov, V. Schneider, E. Shekhtman, K. Sirotkin, D. Slotta, H. Zhang, G. A. McVean, R. M. Durbin, S. Balasubramaniam, J. Burton, P. Danecek, T.

M. Keane, A. Kolb-Kokocinski, S. McCarthy, J. Stalker, M. Quail, J. P. Schmidt, C. J. Davies, J. Gollub, T. Webster, B. Wong, Y. Zhan, A. Auton, C. L. Campbell, Y. Kong, A. Marcketta, R. A. Gibbs, F. Yu, L. Antunes, M. Bainbridge, D. Muzny, A. Sabo, Z. Huang, J. Wang, L. J. M. Coin, L. Fang, X. Guo, X. Jin, G. Li, Q. Li, Y. Li, Z. Li, H. Lin, B. Liu, R. Luo, H. Shao, Y. Xie, C. Ye, C. Yu, F. Zhang, H. Zheng, H. Zhu, C. Alkan, E. Dal, F. Kahveci, G. T. Marth, E. P. Garrison, D. Kural, W.-P. Lee, W. Fung Leong, M. Stromberg, A. N. Ward, J. Wu, M. Zhang, M. J. Daly, M. A. DePristo, R. E. Handsaker, D. M. Altshuler, E. Banks, G. Bhatia, G. del Angel, S. B. Gabriel, G. Genovese, N. Gupta, H. Li, S. Kashin, E. S. Lander, S. A. McCarroll, J. C. Nemesh, R. E. Poplin, S. C. Yoon, J. Lihm, V. Makarov, A. G. Clark, S. Gottipati, A. Keinan, J. L. Rodriguez-Flores, J. O. Korbel, T. Rausch, M. H. Fritz, A. M. Stütz, P. Flicek, K. Beal, L. Clarke, A. Datta, J. Herrero, W. M. McLaren, G. R. S. Ritchie, R. E. Smith, D. Zerbino, X. Zheng-Bradley, P. C. Sabeti, I. Shlyakhter, S. F. Schaffner, J. Vitti, D. N. Cooper, E. V. Ball, P. D. Stenson, D. R. Bentley, B. Barnes, M. Bauer, R. Keira Cheetham, A. Cox, M. Eberle, S. Humphray, S. Kahn, L. Murray, J. Peden, R. Shaw, E. E. Kenny, M. A. Batzer, M. K. Konkel, J. A. Walker, D. G. MacArthur, M. Lek, R. Sudbrak, V. S. Amstislavskiy, R. Herwig, E. R. Mardis, L. Ding, D. C. Koboldt, D. Larson, K. Ye, S. Gravel, The 1000 Genomes Project Consortium, Corresponding authors, Steering committee, Production group, Baylor College of Medicine, BGI-Shenzhen, Broad Institute of MIT and Harvard, Coriell Institute for Medical Research, E. B. I. European Molecular Biology Laboratory, Illumina, Max Planck Institute for Molecular Genetics, McDonnell Genome Institute at Washington University, US National Institutes of Health, University of Oxford, Wellcome Trust Sanger Institute, Analysis group, Affymetrix, Albert Einstein College of Medicine, Bilkent University, Boston College, Cold Spring Harbor Laboratory, Cornell University, European Molecular Biology Laboratory, Harvard University, Human Gene Mutation Database, Icahn School of Medicine at Mount Sinai, Louisiana State University, Massachusetts General Hospital, McGill University, N. National Eye Institute, A global reference for human genetic variation. *Nature*. **526**, 68–74 (2015).
12. J. Neyman, E. S. Pearson, On the Use and Interpretation of Certain Test Criteria for Purposes of Statistical Inference: Part I. *Biometrika*. **20A**, 175–240 (1928).
13. O. J. Dunn, Multiple Comparisons among Means. *Journal of the American Statistical Association*. **56**, 52–64 (1961).
14. T. M. Therneau, T. L. (original S.->R port and R. maintainer until 2009), A. Elizabeth, C. Cynthia, survival: Survival Analysis (2023), (available at https://cran.r-project.org/web/packages/survival/index.html).
15. B. V. Calster, D. Nieboer, Y. Vergouwe, B. D. Cock, M. J. Pencina, E. W. Steyerberg, A calibration hierarchy for risk models was defined: from utopia to empirical data. *Journal of Clinical Epidemiology*. **74**, 167–176 (2016).
16. G. S. Collins, E. O. Ogundimu, D. G. Altman, Sample size considerations for the external validation of a multivariable prognostic model: a resampling study. *Stat Med*. **35**, 214–226 (2016).

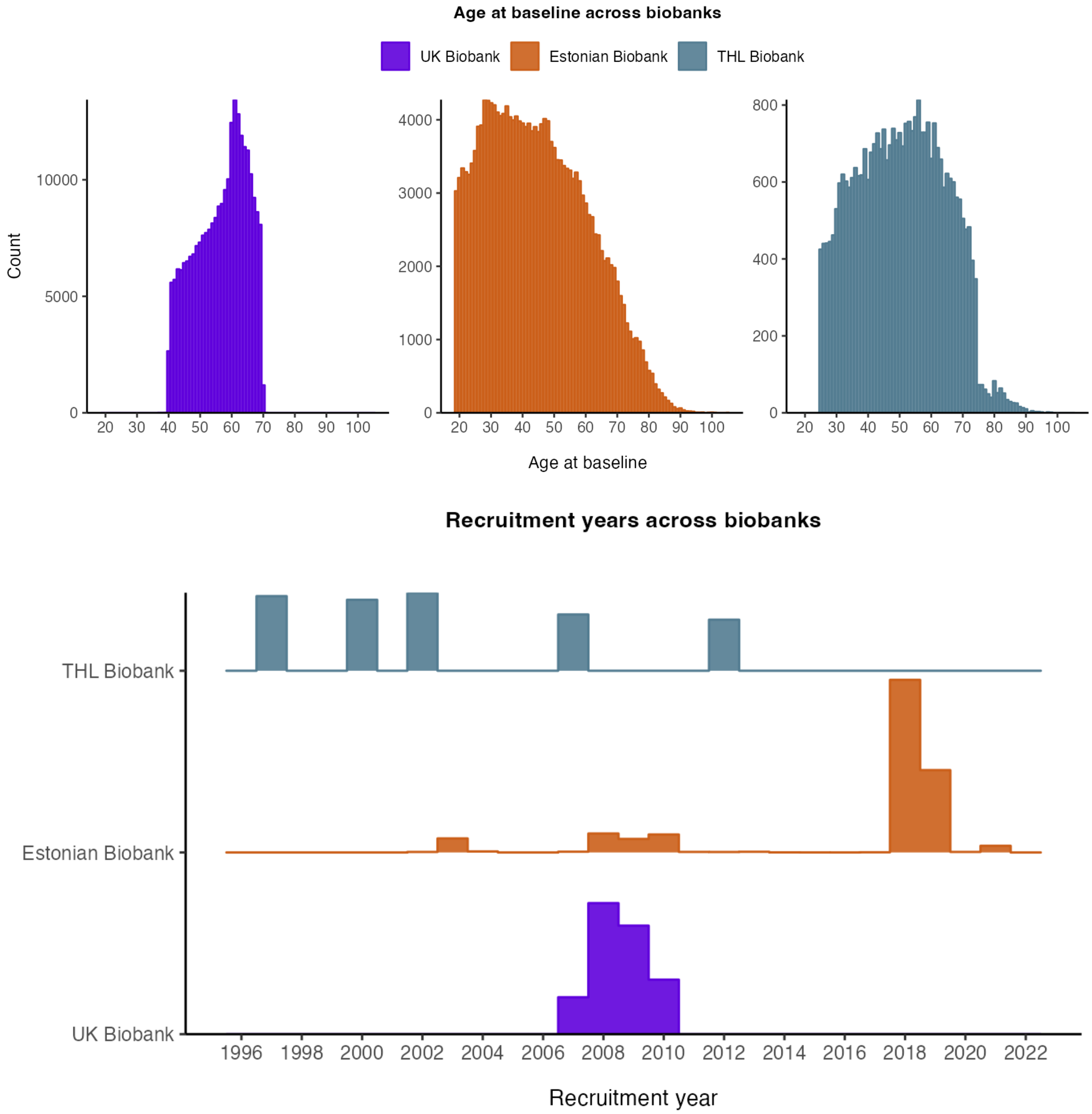


**Fig. S1. Age and recruitment in the three biobanks.**

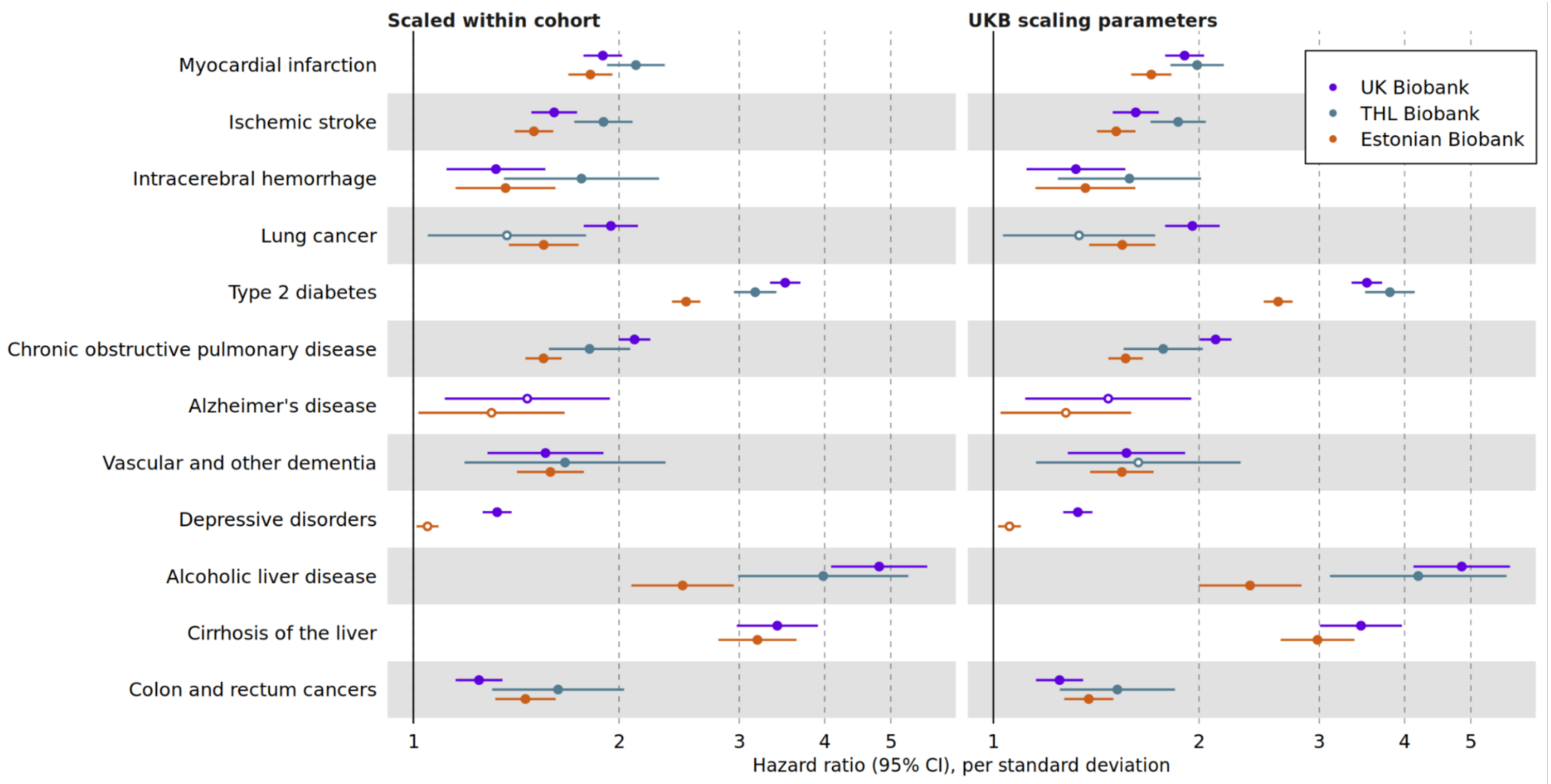


**Fig. S2. Performance of metabolomic scores when scaling within cohort or using scaling parameters from UK Biobank training data.** Hazard ratios are estimated over 4 years of follow-up. Horizontal error bars denote 95% confidence intervals. Closed circles denote p-value < 0.05/12 and open circles p-value >0.05/12.

**A**

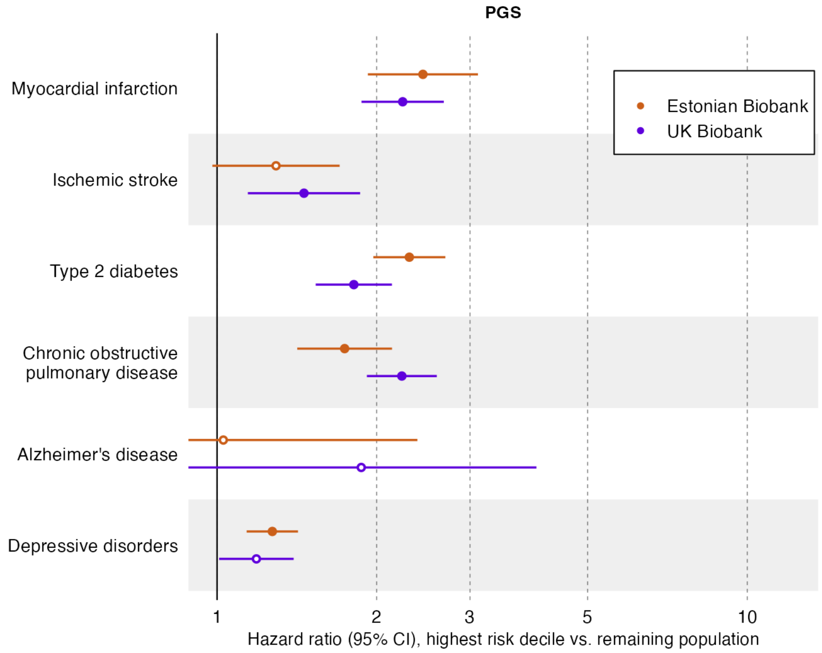


**B**

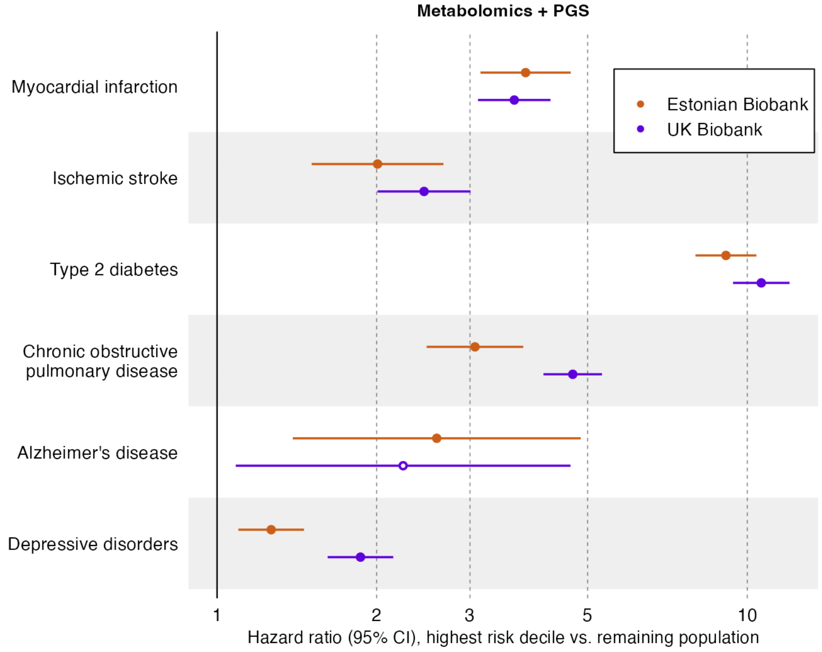


**Fig. S3. Performance of PGS in Estonian Biobank and UK Biobank with 4 years of follow-up.** Horizontal error bars denote 95% confidence intervals. Closed estimate circles denote p-value < 0.05/12 and open circles p-value >0.05/12.

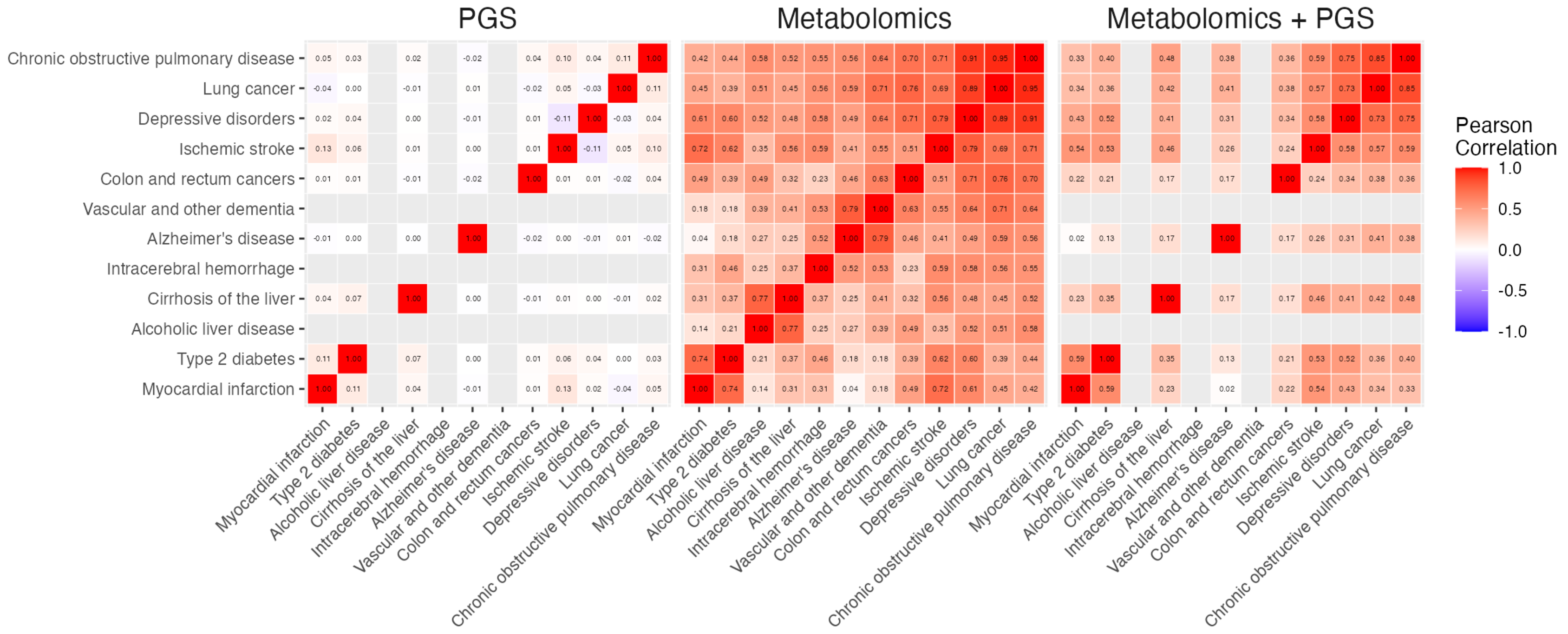


**Fig. S4. Correlations of risk scores between diseases.** Metabolomics matrix is clustered using unweighted pair group method with arithmetic mean. PGS and Metabolomics + PGS matrices are show in the same order as Metabolomics matrix. PGS for Alcoholic liver disease, Intracerebral hemorrhage, and Vascular dementia are missing.

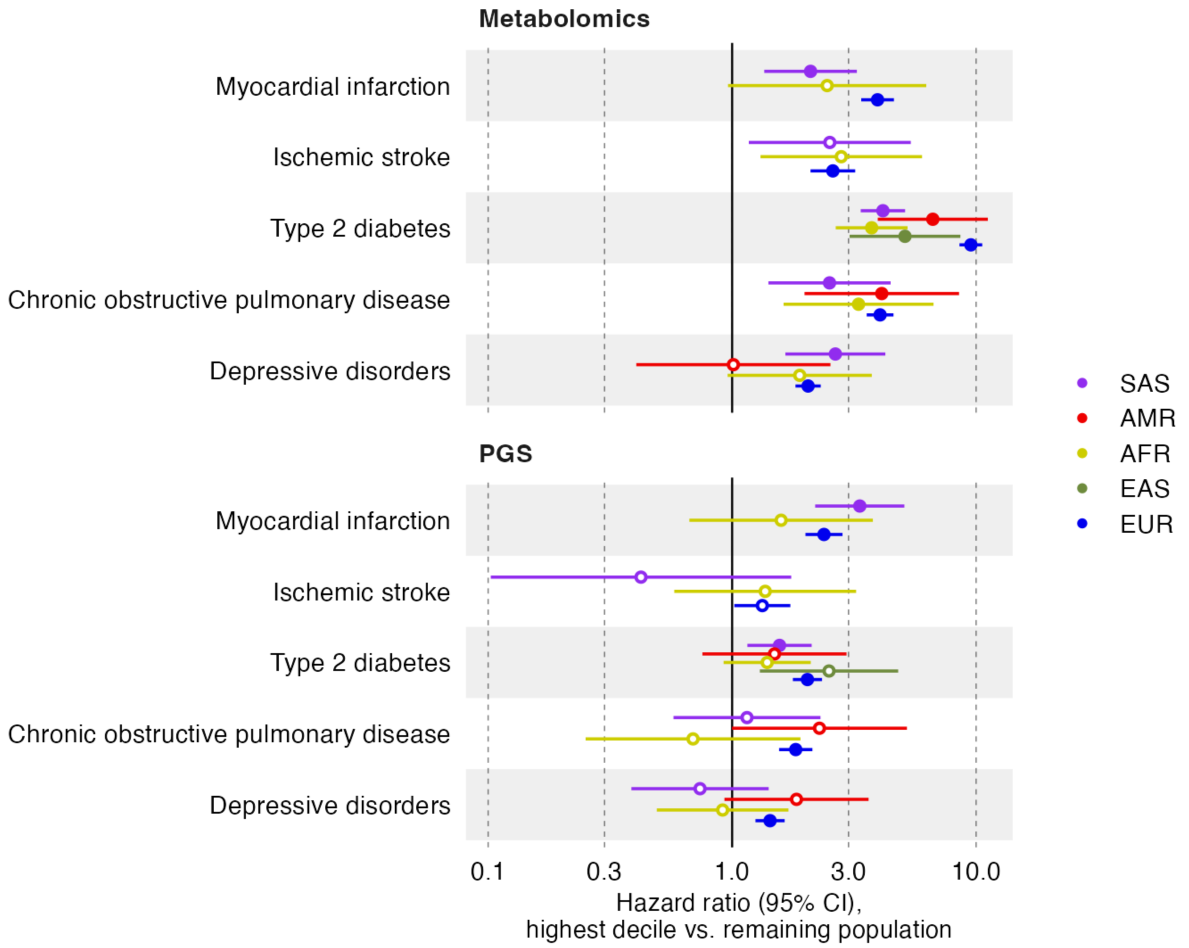


**Fig. S5. Performance of metabolomic score and PGS across ancestries.** Horizontal error bars denote 95% confidence intervals. Closed estimate circles denote p-value <0.05/12 and open circles p-value >0.05/12. AFR = African, AMR = Admixed American, EAS = East Asian, EUR = European, and SAS = South Asian ancestry.

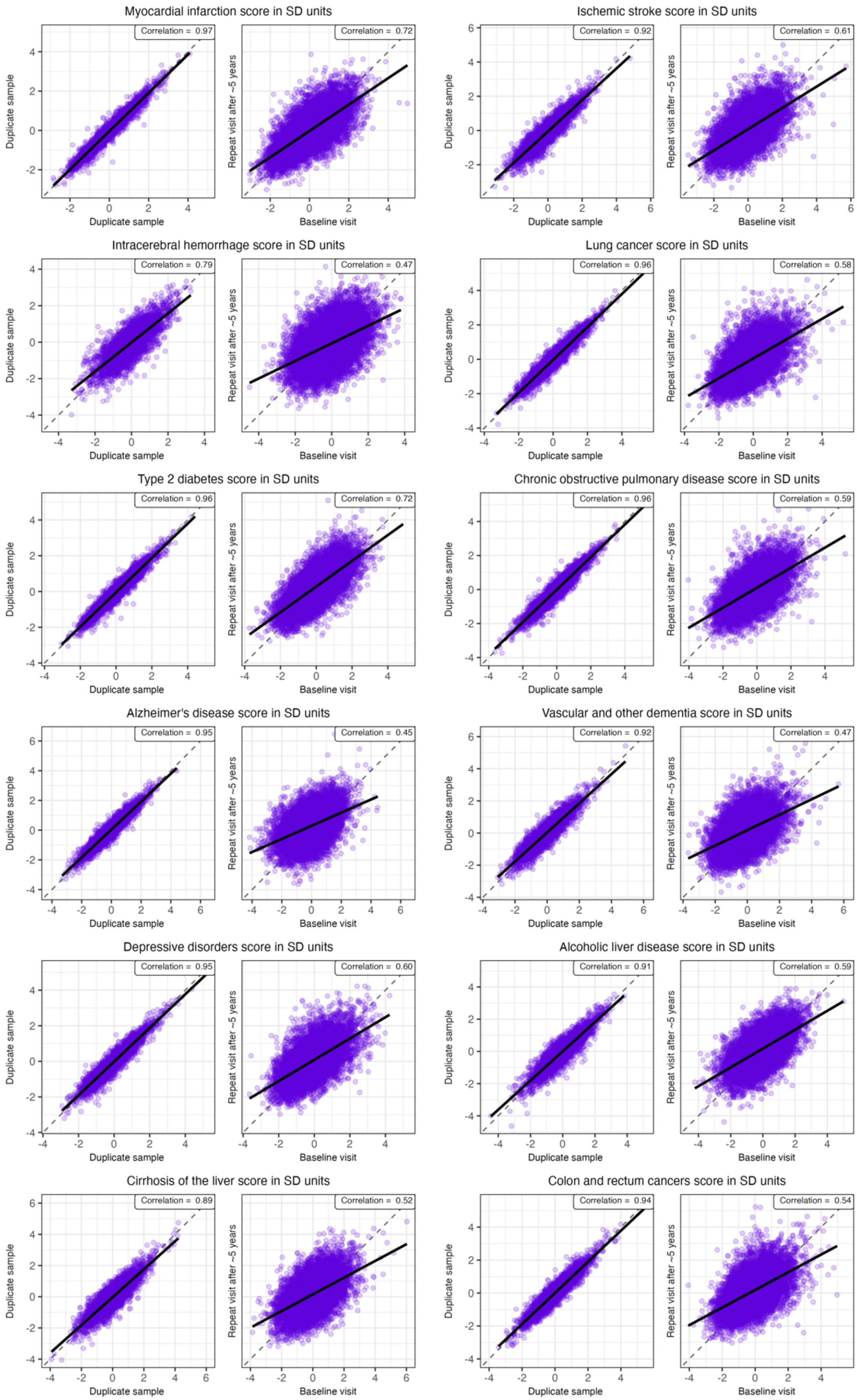
Myocardial infarction score in SD units
Correlation = 0.97
Correlation = 0.72
Ischemic stroke score in SD units
Correlation = 0.92
Correlation = 0.61
Intracerebral hemorrhage score in SD units
Correlation = 0.79
Correlation = 0.47
Lung cancer score in SD units
Correlation = 0.96
Correlation = 0.58
Type 2 diabetes score in SD units
Correlation = 0.96
Correlation = 0.72
Chronic obstructive pulmonary disease score in SD units
Correlation = 0.96
Correlation = 0.59
Alzheimer's disease score in SD units
Correlation = 0.95
Correlation = 0.45
Vascular and other dementia score in SD units
Correlation = 0.92
Correlation = 0.47
Depressive disorders score in SD units
Correlation = 0.95
Correlation = 0.60
Alcoholic liver disease score in SD units
Correlation = 0.91
Correlation = 0.59
Cirrhosis of the liver score in SD units
Correlation = 0.89
Correlation = 0.52
Colon and rectum cancers score in SD units
Correlation = 0.94
Correlation = 0.54
Duplicate sample
Repeat visit after ~5 years
Baseline visit

**Fig S6. Correlations of metabolomic scores between duplicate measurements, and baseline and repeat visit after five years.** Black solid line denotes linear regression line. Dashed line denotes the diagonal.

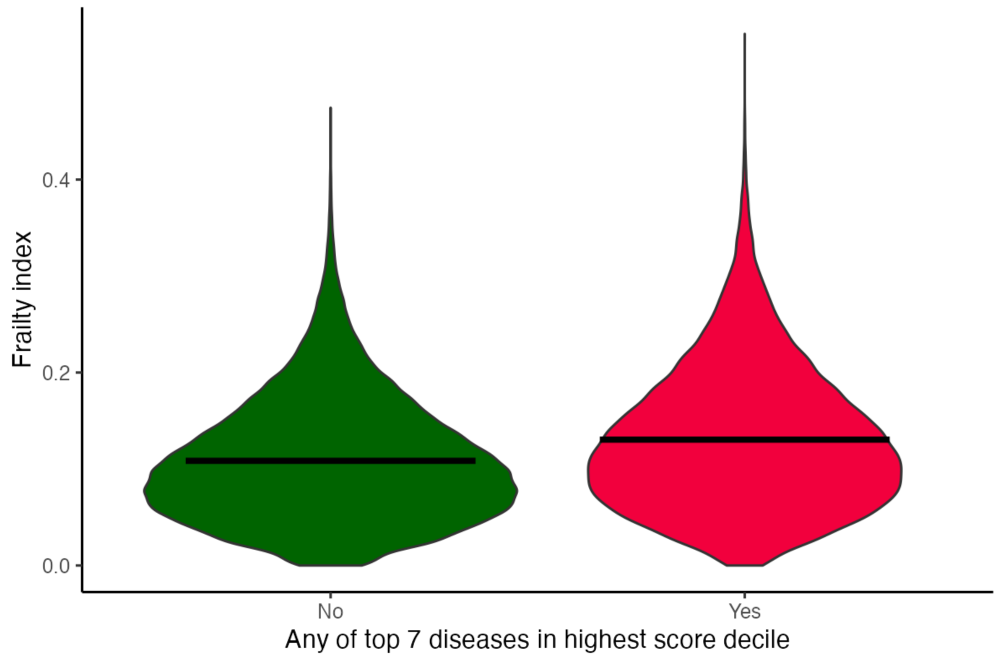


**Fig S7**. Distributions of frailty index between individuals with low and high metabolomic risk. Horizontal lined denote mean frailty indices.

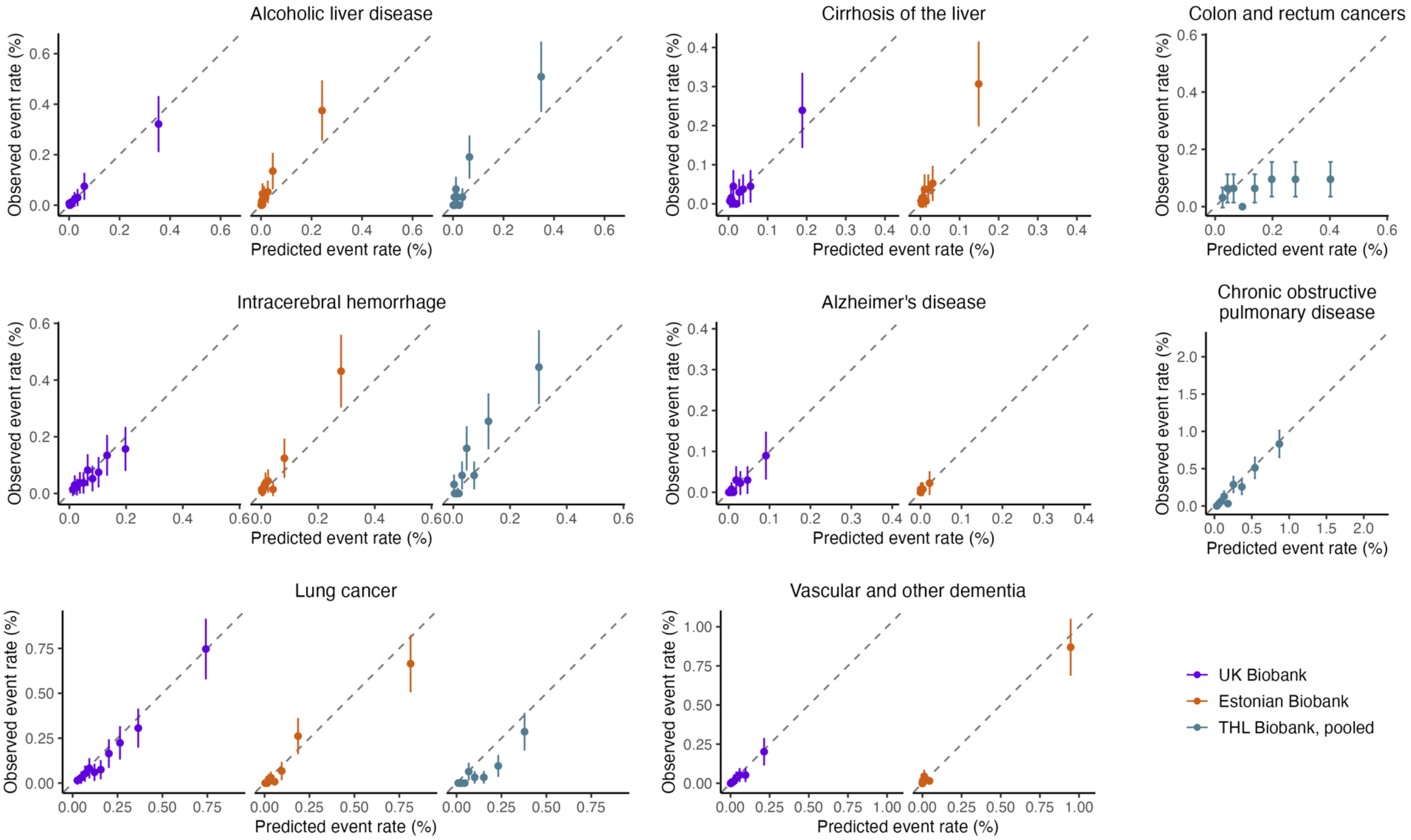


**Fig. S8. Model calibration for diseases and cohorts where number of events < 200.** For each disease outcome, the calibration of three-year observed event rates are shown by 10 equally sized deciles of absolute predicted risk. Vertical lines represent 95% confidence intervals.

| | FINRISK 1997 | HEALTH 2000 | FINRISK 2002 | FINRISK 2007 | FINRISK 2012 |
|---|---|---|---|---|---|
| Number of participants | 7199 | 6847 | 7527 | 5430 | 4925 |
| Age at blood sample (median, [IQR]) | 48.0 [37.0-59.0] | 53.0 [42.0-65.0] | 48.0 [37.0-58.0] | 52.0 [40.0-63.0] | 53.0 [40.0-64.0] |
| Females (%) | 50.7 | 55.3 | 55.5 | 53.4 | 52.1 |
| Body mass index (kg/m^2, median, [IQR]) | 26.0 [23.4-28.9] | 26.4 [23.7-29.5] | 26.2 [23.5-29.2] | 26.3 [23.7-29.6] | 26.3 [23.7-29.6] |
| Smoking prevalence (current, %) | 34.7 | 39.1 | 37.7 | 30.9 | 28.8 |
| Cholesterol lowering medication (%) | 4.6 | 7.4 | 9.1 | 16.5 | 18.5 |
| Follow-up time (median, [IQR]) | 18.8 [18.8-18.9] | 15.1 [14.9-15.2] | 13.8 [13.8-13.9] | 8.9 [8.8-8.9] | 3.8 [3.8-3.9] |
| Recruitment period | 1997 | 2000 | 2002 | 2007 | 2012 |

**Table S1. Epidemiological characteristics of THL Biobank cohorts.**

| Disease | HR per SD | 95% CI | P-value | N | N cases |
|---|---|---|---|---|---|
| Alzheimer's disease | 0.95 | (0.87, 1.04) | 2.56E-01 | 135,023 | 440 |
| Colon and rectum cancers | 1.00 | (0.95, 1.05) | 9.09E-01 | 134,267 | 1,559 |
| Chronic obstructive pulmonary disease | 1.00 | (0.97, 1.03) | 8.35E-01 | 132,509 | 3,756 |
| Depressive disorders | 0.98 | (0.95, 1.01) | 1.71E-01 | 123,183 | 3,963 |
| Type 2 diabetes | 0.92 | (0.90, 0.95) | 3.93E-10 | 129,269 | 4,143 |
| Cirrhosis of the liver | 1.01 | (0.94, 1.09) | 7.19E-01 | 134,859 | 337 |
| Lung cancer | 1.01 | (0.96, 1.06) | 7.97E-01 | 134,867 | 1,104 |
| Myocardial infarction | 0.99 | (0.95, 1.03) | 5.94E-01 | 131,936 | 2,319 |
| Ischemic stroke | 1.01 | (0.96, 1.05) | 7.94E-01 | 133,090 | 1,708 |

**Table S4. Estimates for interaction between metabolomic and polygenic scores.** Estimates are shown for interaction term `metabolic score * PGS`. HR = Hazard Ratio. SD = Standard deviation, CI = Confidence interval, N = Total number of samples, N cases = Number of cases within samples.

| Metabolomic score | Correlation between baseline and follow-up visit | Correlation between duplicate measurements |
|---|---|---|
| Alcoholic liver disease | 0.59 | 0.91 |
| Alzheimer's disease | 0.45 | 0.95 |
| Colon and rectum cancers | 0.54 | 0.94 |
| Chronic obstructive pulmonary disease | 0.59 | 0.96 |
| Depressive disorders | 0.60 | 0.95 |
| Diabetes | 0.72 | 0.96 |
| Cirrhosis of the liver | 0.52 | 0.89 |
| Intracerebral hemorrhage | 0.47 | 0.79 |
| Lung cancer | 0.58 | 0.96 |
| Myocardial infarction | 0.72 | 0.97 |
| Ischemic stroke | 0.61 | 0.92 |
| Vascular and other dementia | 0.47 | 0.92 |

**Table S5. Correlations of metabolomic scores between baseline and repeat visit after 5 years.**

| Variable | Low risk group mean (SD) or N [%] | High risk group mean (SD) or N [%] | Difference in mean (95% CI) | p-value |
|---|---|---|---|---|
| Frailty index | 0.11 (0.06) | 0.13 (0.07) | 0.02 (0.02 - 0.02) | < 2.2e-16 |
| Current smoking | 6424 [7.35] | 5555 [15.95] | -- | < 2.2e-16 |
| BMI (kg/m2) | 26.41 (4.01) | 28.9 (5.3) | 2.49 (2.43 - 2.55) | < 2.2e-16 |
| Systolic blood pressure (mmHg) | 136.29 (18.4) | 140.94 (18.77) | 4.65 (4.41 - 4.89) | 1.7e-319 |
| Total cholesterol (mmol/l) | 5.78 (1.01) | 5.78 (1.24) | 0 (-0.02 - 0.01) | 0.77 |
| HDL cholesterol (mmol/l) | 1.52 (0.35) | 1.35 (0.42) | -0.17 (-0.17 - -0.16) | < 2.2e-16 |
| Waist circumference (cm) | 87.15 (11.85) | 94.15 (13.97) | 6.99 (6.83 - 7.16) | < 2.2e-16 |
| Hip circumference (cm) | 101.91 (7.98) | 105.47 (10.32) | 3.56 (3.44 - 3.68) | < 2.2e-16 |
| Age (years) | 55.9 (8.04) | 56.89 (8.07) | 0.99 (0.89 - 1.09) | 2.3e-83 |
| Male sex | 37664 [42.94] | 16869 [48.17] | -- | 2.7e-62 |
| Statin use | 9372 [10.68] | 5580 [15.93] | -- | 3.8e-142 |

**Table S6. Clinical characteristics between individuals with low and high metabolomic risk score.** Low risk group includes individuals whose metabolomic risk is not in the highest decile for any of the 7 scores considered. High risk group includes individuals who has at least one of the 7 metabolic risks in the highest decile. SD = standard deviation. CI = Confidence interval.
